# Surveillance colonoscopy in patients with quiescent inflammatory bowel disease is associated with increased post-procedure steroid prescriptions: a national database study

**DOI:** 10.1101/2025.08.05.25332988

**Authors:** Derek Liu, Chiraag Kulkarni, Gavin Hui, C. William Pike, Carolina Tropini, Saurabh Gombar, Sidhartha R. Sinha

## Abstract

Surveillance colonoscopy reduces colorectal cancer (CRC) risk in inflammatory bowel disease (IBD), but anecdotal evidence suggests a link between surveillance colonoscopy and symptoms flare, which may affect patient compliance with CRC surveillance. We investigated the effects of surveillance colonoscopy in quiescent IBD using a national database to conduct a retrospective, propensity score-matched (PSM) cohort study of 1,717 patients with IBD and 1,717 patients without IBD who underwent colonoscopy. Strict inclusion criteria were used to select patients with quiescent IBD undergoing surveillance colonoscopy. We demonstrated that patients with quiescent IBD prior to colonoscopy received significantly more post-colonoscopy steroid prescriptions compared to matched controls (OR 1.46 [1.20, 1.77], p<0.001). Steroid prescriptions were more likely in patients with IBD at longer follow-up intervals, suggesting a delayed mechanism of onset for post-colonoscopy inflammation. No significant differences between groups were observed for the other outcomes of post-procedure emergency room and hospital visits or enteric infections. Our results suggest a potential risk of symptom exacerbation, evidenced by increased steroid prescriptions, following surveillance colonoscopy. Given that nearly 25% of patients with IBD do not receive surveillance colonoscopy at the recommended intervals, active post-procedure symptoms may be a contributing factor that warrants further investigation.

## 1. Introduction

Inflammatory bowel disease (IBD), including Crohn’s disease (CD) and ulcerative colitis (UC), is a chronic inflammatory disorder of the gastrointestinal tract that increases the risk of high-grade dysplasia and colorectal cancer (CRC). IBD is most commonly diagnosed between the ages of 20 to 40, and the cumulative risk of developing CRC in patients with IBD has been estimated to be 1%, 3%, and 7% at 10, 20, and 30 years of disease, respectively [1,2]. In comparison, the 10-year cumulative risk for the average-risk population at age 50 years has been estimated to be 0.4%, and the global cumulative risk of colorectal cancer between 0-74 years has been shown to be about 2% [3-5]. IBD is a significant risk factor in CRC development and endoscopic surveillance is widely recommended by gastroenterology societies, typically starting 8 years after diagnosis [6-10].

It has previously been suggested that bowel preparation for colonoscopy or the procedure itself may exacerbate IBD symptoms. A small, single-center prospective study in patients with UC found that 6 out of 51 patients had clinical relapse following colonoscopy defined by a score of 5 or greater on the Simple Colitis Clinical Activity Index (SCCAI) during the week after colonoscopy; patients with a higher baseline SCCAI prior to colonoscopy were more likely to have an increased post-procedure SCCAI [11]. A separate study involving 109 patients with inactive UC undergoing surveillance colonoscopy found that over 23% experienced increased SCCAI scores within four weeks, prompting escalated medication regimens in a subset of these patients [12]. A subsequent retrospective self-controlled cohort study suggested that there is an increased risk of emergency room (ER) visits following colonoscopy for any indication in patients with IBD [13]. However, this study was likely limited by inclusion of patients with active IBD disease given the liberal inclusion criteria, a study population with limited diversity, and a very high, non-representative, percentage of patients (>90%) with IBD not on maintenance therapy, which would likely impact propensity to have clinical symptoms [14].

To our knowledge, there have not been large cohort studies performed to investigate the relationship between colonoscopy and adverse GI symptom effects in patients with quiescent IBD. Thus, the risk of GI symptom exacerbation following surveillance colonoscopy as well as modulators of this risk remain incompletely characterized.

To this end, we conducted a large retrospective cohort study with propensity score-matched (PSM) analysis to evaluate the risk of GI symptom exacerbation in patients with quiescent IBD following surveillance colonoscopy.

## 2. Results

### 2.1 Cohort demographics

Our cohort prior to propensity score matching consisted of 26,825 patients, including 1,825 patients with IBD and 25,000 patients without IBD who underwent colonoscopy between 2010 and 2024. The final propensity score-matched cohort consisted of 1,717 patients with IBD and 1,717 matched controls. In the matched cohort, 50.3% of patients with IBD and 49.3% of patients without IBD were female. Mean age for the IBD group was 58.7 years old, while mean age for the non-IBD group was 59.1 years old. Demographic data and descriptive statistics for the cohort are presented in Table 1.

**Table 1.** Demographic data of propensity score-matched cohort. Data are expressed as counts with corresponding percentage or as mean/score with standard deviation. IBD - inflammatory bowel disease; sd - standard deviation.

|  | **IBD group** | **non-IBD group** |
| --- | --- | --- |
| **N** | 1,717 | 1,717 |
| **Sex (%)** |  |  |
| Female | 863 (50.3%) | 846 (49.3%) |
| Male | 854 (49.7%) | 871 (50.7%) |
| **Mean age (sd)** | 58.7 (12.7) | 59.1 (12.3) |
| **Age (%)** |  |  |
| <18 yr | 0 (0%) | 0 (0%) |
| 18-29 yr | 29 (1.7%) | 55 (3.2%) |
| 30-39 yr | 147 (8.6%) | 79 (4.6%) |
| 40-49 yr | 243 (14.2%) | 199 (11.6%) |
| 50-59 yr | 411 (23.9%) | 452 (26.3%) |
| 60-69 yr | 506 (29.5%) | 595 (34.7%) |
| 70-79 yr | 381 (22.2%) | 337 (19.6%) |
| >=80 yr | 0 (0%) | 0 (0%) |
| **Race (%)** |  |  |
| White | 743 (43.3%) | 696 (40.5%) |
| Black | 63 (3.7%) | 52 (3.0%) |
| Asian | 17 (1.0%) | 29 (1.7%) |
| Other | 894 (52.1%) | 940 (54.7%) |
| **Hispanic (%)** | 49 (2.9%) | 86 (5.0%) |
| **Charlson comorbidity score (sd)** | 3.3 (3.2) | 3.5 (3.4) |

### 2.2 Validation of electronic phenotype for quiescent IBD

Using the Stanford Research Repository (STARR), a cohort of 100 patients with IBD was randomly selected from patients who matched our inclusion and exclusion criteria for quiescent disease. Chart review of endoscopic reports was performed. We found that only 6 patients (6%) in this independent cohort, who would have met inclusion criteria for the original study design, had significant inflammation (Mayo Score >1 or SES-CD score >6) at time of colonoscopy.

### 2.3 Primary and secondary outcomes

In propensity score-matched analysis, the primary outcome of composite emergency room visits and hospitalizations of any cause during days 7 to 30 post-colonoscopy was not significantly different between patients with IBD and the control group (PSM OR 0.778, 95% CI 0.536-1.13, p=0.19) (Figure 1, Table 2). Patients with IBD did receive steroid prescriptions following colonoscopy at a significantly higher rate than the control group, with 16% of patients with IBD receiving steroids compared to 11% of matched controls (PSM OR 1.46, 95% CI 1.20-1.77, p<0.001) (Figure 1, Table 2). Enteric infections were not significantly different between the IBD and control groups (PSM OR 0.299, 95% CI 0.0528-1.16, p=0.092) (Figure 1, Table 2).

**Table 2.** Propensity score-matched results of primary and secondary outcomes. IBD - inflammatory bowel disease; OR - odds ratio; CI - confidence interval.

| **Composite emergency room visits and hospitalizations** |  |  |  |  |
| --- | --- | --- | --- | --- |
| **Comparison** | **IBD group (%)** | **Control group (%)** | **OR (95% CI)** | **p-value** |
| IBD vs control | 51/1,717 (3.0%) | 65/1,717 (3.8%) | 0.778 (0.536, 1.13) | 0.19 |
| **Steroid prescriptions** |  |  |  |  |
| **Comparison** | **IBD group (%)** | **Control group (%)** | **OR (95% CI)** | **p-value** |
| IBD vs control | 270/1,717 (16%) | 195/1,717 (11%) | 1.46 (1.20, 1.77) | <0.001 |
| **Enteric infections** |  |  |  |  |
| **Comparison** | **IBD group (%)** | **Control group (%)** | **OR (95% CI)** | **p-value** |
| IBD vs control | 3/1,717 (0.17%) | 10/1,717 (0.58%) | 0.299 (0.0528, 1.16) | 0.092 |

**Figure 1.**
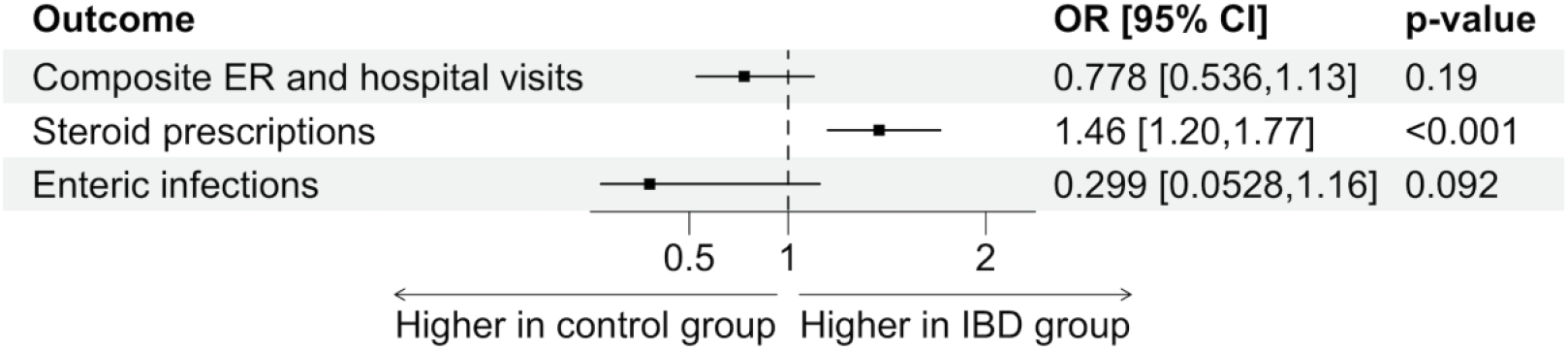
Propensity score-matched analysis of primary outcome (composite ER and hospital visits) and secondary outcomes (steroid prescriptions, enteric infections) following colonoscopy in patients with quiescent IBD and matched controls. ER - emergency room; OR - odds ratio; CI - confidence interval.

### 2.4 Sensitivity analyses

When stratified by IBD disease type, patients with CD (PSM OR 1.41, 95% CI 1.07-1.88, p=0.016) and UC (PSM OR 1.66, 95% CI 1.28-2.16, p<0.001) both received significantly more steroid prescriptions following colonoscopy compared to propensity score-matched controls (Figure 2, Table 3). Both subsets of patients did not have significantly different composite emergency room visits and hospitalizations compared to patients without IBD.

**Table 3.**
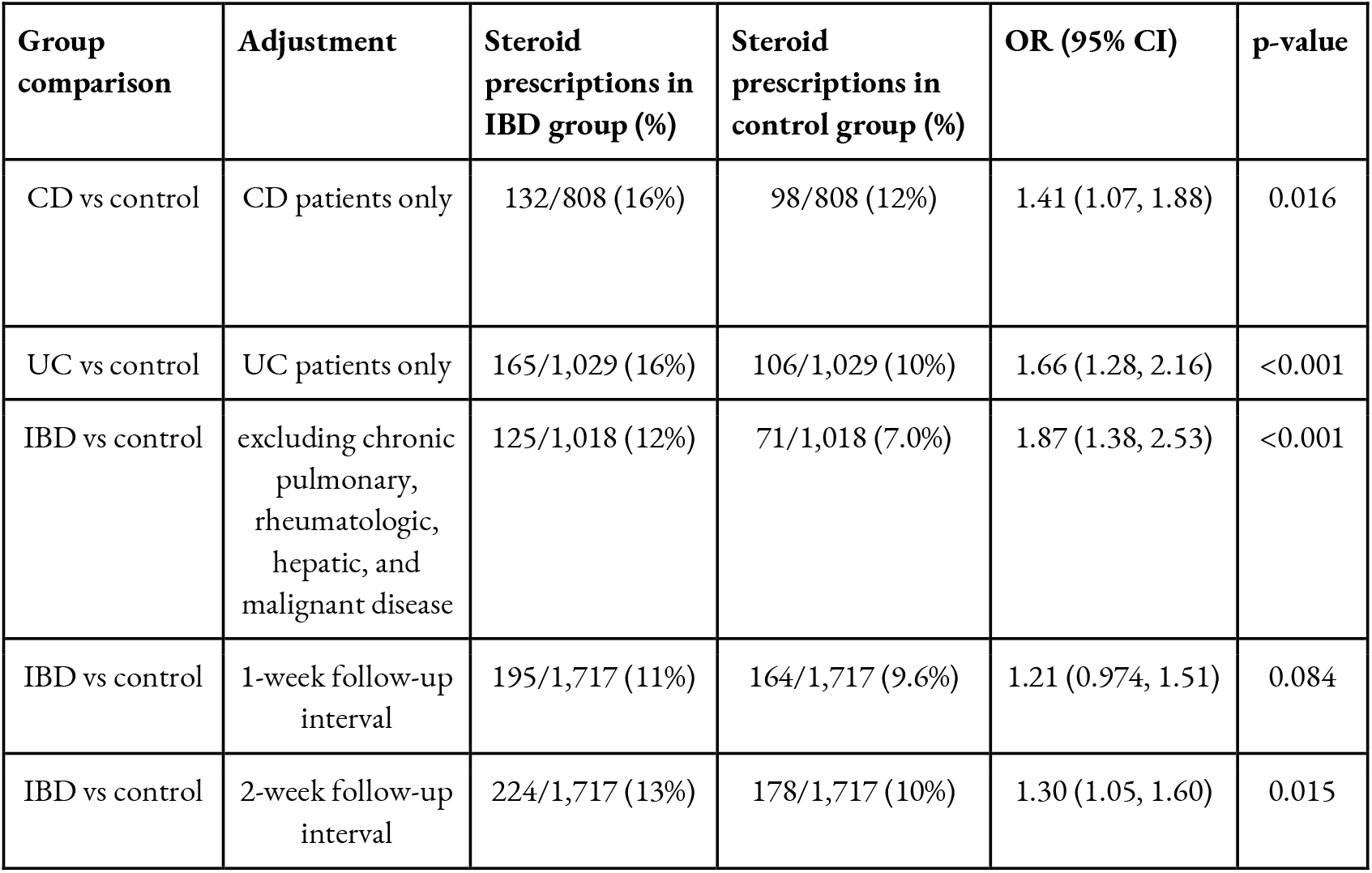
Propensity score-matched results of sensitivity analyses. CD - Crohn’s disease; UC - ulcerative colitis; IBD - inflammatory bowel disease; OR - odds ratio; CI - confidence interval.

**Figure 2.**
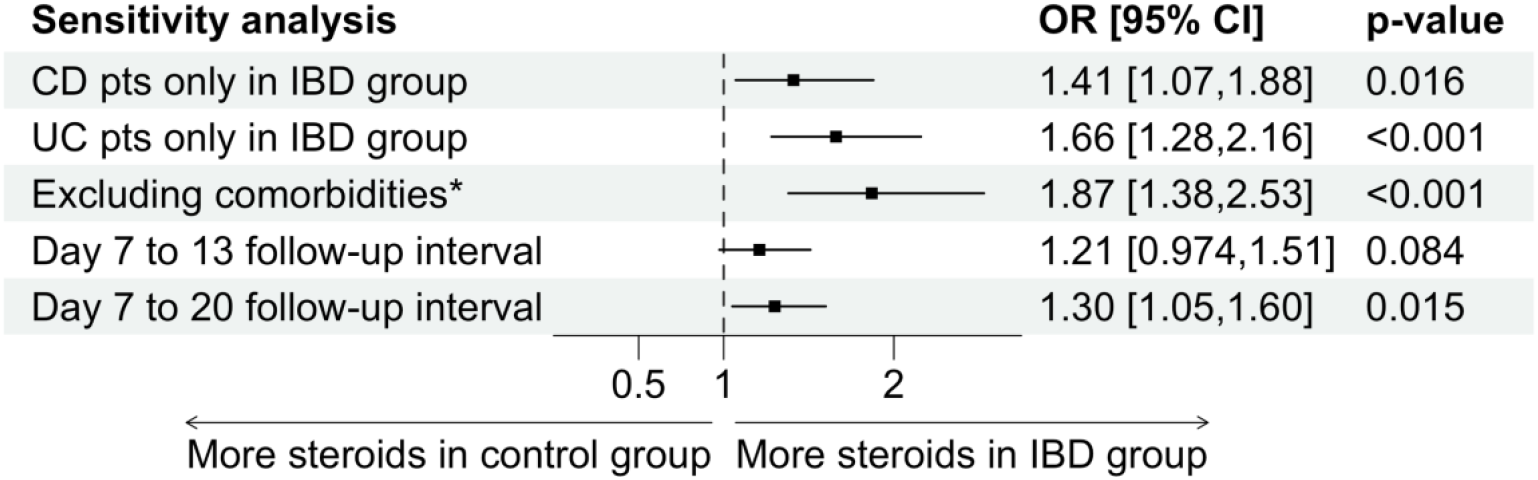
Propensity score-matched results of sensitivity analyses. Post-colonoscopy steroid prescriptions were assessed in patients with quiescent IBD compared to matched controls after stratification by IBD type, exclusion of comorbidities that may predispose to steroid treatment, and changes in follow-up interval duration. ^*^Chronic pulmonary, rheumatologic, hepatic, and malignant comorbidities; CD - Crohn’s disease; UC - ulcerative colitis; IBD - inflammatory bowel disease; OR - odds ratio; CI - confidence interval.

Steroid prescription rates were evaluated after excluding patients with comorbidities that may predispose to receiving steroid treatment (Figure 2, Table 3). When patients with chronic pulmonary, rheumatologic, hepatic, and cancer comorbidities were excluded from the study cohort, the rate of steroid prescriptions following colonoscopy remained significantly higher in the IBD group compared to propensity score-matched controls (12% vs 7.0%, PSM OR 1.87, 95% CI 1.38-2.53, p<0.001).

Steroid prescription rates were also assessed at different follow-up time intervals after colonoscopy (Figure 2, Table 3). During a one-week interval (days 7-13) post-colonoscopy, there was no significant difference in steroid prescriptions between patients with IBD and the control group (PSM OR 1.21, 95% CI 0.974-1.51, p=0.084). When assessed over a two-week interval (days 7-20) following colonoscopy, however, there was a significantly higher rate of steroid prescriptions in patients with IBD compared to the control group (PSM OR 1.30, 95% CI 1.05-1.60, p=0.015).

## 3. Discussion

In this study, we found that patients with quiescent IBD received more steroid prescriptions following surveillance colonoscopy compared to propensity score-matched controls, though they did not have different rates of post-procedure composite ER visits and hospitalizations. A prior large cohort, self-controlled study found that patients with IBD had an increased risk of ER visits during the immediate period following colonoscopy compared to a control interval further removed from colonoscopy [13]. However, that study was likely limited by including patients with IBD who underwent colonoscopy for any indication and thereby including patients with active disease flares, lack of diversity in the patient population, and a vast majority (>90%) of patients not on any IBD maintenance therapies. Overall, our findings suggest that patients with IBD may be at higher risk of developing GI symptom exacerbation post-colonoscopy. It is likely that these symptoms are mild to moderate in nature, warranting steroid prescription but not ER visits or hospitalizations.

Our findings were robust, with consistent findings across multiple sensitivity analyses. Patients with both CD and UC received significantly more steroid prescriptions post-colonoscopy compared to matched controls. Furthermore, when patients with chronic pulmonary, rheumatologic, hepatic, and malignant comorbidities (which may predispose to steroid treatment) were excluded from analysis, the rate of post-colonoscopy steroid prescriptions was significantly increased in the IBD group by 87% compared to matched controls.

Of note, the steroid prescriptions captured in our study were for any indication. The initial unmatched control group appeared to be representative of the wider domestic population as 7.7% received steroid prescriptions, which is similar to a recent estimate of 6.8% oral corticosteroid use in the United States with a range of 2.2% to 17.5% in other countries [16]. It is possible that the higher rate of steroid prescriptions observed in the IBD group compared to the control group may have been driven by non-IBD-related reasons for steroid prescriptions. However, we attempted to minimize these non-IBD-related confounders through propensity score matching and sensitivity analysis. Notably, after propensity score matching, Charlson comorbidity score was similar between the IBD and control groups (3.3 vs 3.5, respectively). Furthermore, we observed increased steroid prescriptions in the IBD group compared to the control group after excluding patients with pulmonary, rheumatologic, hepatic, and malignant comorbidities that would predispose patients to other causes of steroid prescriptions.

The mechanism for colonoscopy-related symptom exacerbation in patients with IBD remains uncertain though several hypotheses have been proposed. Our data showed that steroids were more likely to be prescribed in patients with IBD during the 4-week and 2-week follow-up intervals compared to the 1-week follow-up interval after colonoscopy, suggesting a delayed mechanism. Previous work has suggested that sodium phosphate-containing bowel preparation agents can lead to inflammation and ulcerative abnormalities [17-19]. A large prospective study of 634 patients showed that preparation-induced mucosal inflammation was 10-fold greater with sodium phosphate (an osmotic laxative) and sodium picosulfate/magnesium citrate (stimulant and osmotic laxatives, respectively) bowel cleansing agents compared to polyethylene glycol (an osmotic laxative) [20]. Recent studies have also shown that bowel preparation has a different effect on the gut microbiota of patients with IBD compared to healthy controls [21,22]. Furthermore, our prior work in animal models showed that polyethylene glycol bowel preparation increases GI osmolality and leads to increased susceptibility to pathogens and microbiota changes [23]. Another recent study showed that bowel preparation and colonoscopy were associated with gut microbiota changes and decreased levels of fecal short-chain fatty acids, which may be related to GI symptoms that developed after colonoscopy [24]. It will be important to further investigate whether certain colonoscopy bowel preparation agents or microbiota changes predispose patients with IBD to symptom exacerbations.

This study has several strengths, including utilizing a large cohort derived from a nationally representative database as well as employing strict inclusion and exclusion criteria to select for patients with quiescent IBD. In addition to requiring patients to meet clinical and lab criteria indicating high likelihood of quiescent disease, we also omitted CPT codes (such as CPT 45367, CPT 45380, etc.) that are often used for diagnostic procedures, which may be performed for active symptoms or inflammation. Our rigorous selection criteria yielded 1,825 patients with IBD, of which 1,717 patients were included in the PS-matched analysis. In order to minimize confounding between the IBD and control group, we used propensity score-matching between the two groups. Additionally, the consistency of results across multiple sensitivity analyses, both in direction and magnitude, further reinforces the reliability and validity of the findings.

Our study also has some limitations. First, we lacked access to endoscopic data in our national cohort, raising the possibility that steroids could be prescribed for pre-existing inflammation visualized at time of endoscopy rather than for GI symptoms that developed post-procedurally. To our knowledge, there is no national database that fully captures endoscopic, histologic, and clinical metadata for IBD. To address this limitation, we used strict criteria to select for patients with quiescent IBD and validated our selection criteria in an external cohort. In the independent validation cohort in STARR, only 6% of patients with IBD who were prescribed steroids had significant inflammation on endoscopy that would warrant steroid treatment. This low percentage of patients, representing patients with pre-existing inflammation at time of colonoscopy, would not account for the increased rate of post-colonoscopy steroid prescriptions found in the IBD group from our primary analysis. Second, our dataset had limited annotation of IBD disease location so the effects of colonoscopy on specific disease phenotypes is limited. Additional large cohorts with comprehensive clinical annotations are needed to further characterize which patients with IBD are at highest risk of developing adverse symptoms following colonoscopy.

In conclusion, patients with quiescent IBD who undergo colonoscopy were more likely to receive steroid prescriptions following their procedure, which is likely driven by GI symptom exacerbation after colonoscopy rather than active IBD at time of colonoscopy. Given up to 25% of patients with IBD do not receive surveillance colonoscopies at the recommended intervals [25], providers should be aware of the risk of post-colonoscopy symptom exacerbation as possibly contributing to lower adherence rates to CRC screening in patients with IBD and serving as a potential barrier to CRC detection and prevention in this high-risk population.

## 4. Methods

### 4.1 Cohort selection

Using the EVERSANA EHR dataset, a large representative national database, we identified all patients who underwent colonoscopy from 2010 to 2024. The database consists of over 120 million de-identified patients in the US. The data are generated at the point of care and entered into multiple electronic medical record (EMR) systems. The data is sourced from >2,000 outpatient/ambulatory health centers, >500 hospitals, >30 health systems (including academic medical centers) and >50 unique EMR platform providers (e.g. EPIC, CERNER, etc.).

Patients with a history of IBD who underwent colonoscopy were identified using International Classification of Diseases (ICD) codes and Current Procedural Terminology (CPT) codes (see Table S1, which lists included codes). In addition to ICD and CPT codes, strict inclusion criteria were used to select for patients with quiescent IBD undergoing surveillance colonoscopy, including requiring at least 8 years of disease history, excluding patients with a calprotectin level greater than 250 mcg/g in the 90-day period preceding colonoscopy, and excluding days 0 to 6 from the post-colonoscopy follow-up interval (a time period where incidentally found active IBD found on colonoscopy would likely be treated). The control group consisted of patients without a diagnosis of IBD who underwent colonoscopy based on any colonoscopy CPT code and had at least 90 days of history prior to colonoscopy. In order to ensure that the control group did not include patients with IBD, patients could not have any ICD coding for IBD or a calprotectin level greater than 250 mcg/g within 90 days prior to colonoscopy. In patients with multiple colonoscopies matching the inclusion criteria, only the first colonoscopy and follow-up period were included in the analyses.

### 4.2 Data analysis

The primary outcome evaluated was a composite measure of emergency room visits and hospitalizations of any cause during a follow-up interval of days 7 to 30 post-colonoscopy. Days 0 to 6 post-colonoscopy were excluded from analyses as some patients with IBD may have significant endoscopic inflammation despite being asymptomatic and with negative inflammatory markers [15]. These patients would likely receive steroids during this immediate post-procedure time interval for the pre-existing inflammation, which is unlikely to be due to the preparation or procedure itself. The secondary outcomes evaluated were steroid prescriptions and enteric infections during days 7 to 30 post-colonoscopy. We conducted sensitivity analyses to assess whether stratification by IBD disease subtype (CD, UC), exclusion of patients with comorbidities predisposing to steroid treatment (pulmonary, rheumatologic, hepatic, and malignant disease), and different post-colonoscopy follow-up time intervals (1-week, 2-week) had a modifying effect on outcomes.

### 4.3 Validation of electronic phenotype for quiescent IBD

The Stanford Research Repository (STARR) was used to generate an independent cohort of patients with IBD for chart review to validate our inclusion and exclusion criteria for quiescent IBD. In STARR, patients with quiescent IBD who received a glucocorticoid prescription between 7 to 30 days following their most recent colonoscopy were identified by querying for the selection criteria specified above. From the patients identified, 100 patients were randomly selected for chart review to assess whether their colonoscopy report noted significant inflammation (Mayo Score >1 or SES-CD score >6) warranting steroid treatment.

### 4.4 Statistical methods

Categorical variables were summarized using absolute counts and percentages. Outcomes were assessed by calculating the odds ratio along with 95% confidence intervals and corresponding p-values. Matching was done 1:1, with caliper distance of 0.2. Distance tolerance to be considered for exact matching was 0.01. Propensity score matching was performed using all components of the Charlson Comorbidity Index (solid tumor, leukemia, lymphoma, diabetes, congestive heart failure (CHF), myocardial infarction (MI), peripheral vascular disease (PVD), chronic obstructive pulmonary disease (COPD), cerebrovascular disease (CVD), dementia, hemiplegia, liver disease, chronic kidney disease (CKD), peptic ulcer disease (PUD), rheumatic disease, and HIV), year of entry into the cohort, medications, age, sex, and race/ethnicity. Standardized mean difference was calculated for each variable. Propensity score distributions were checked to ensure adequate overlap.

### 4.5 Ethical considerations

All methods were carried out in accordance with relevant guidelines and regulations. All experimental protocols were approved by the Institutional Review Board of Stanford University. Informed consent was waived by the Institutional Review Board of Stanford University due to the use of retrospective, de-identified data.

## 5. Acknowledgements

We would like to acknowledge the support of The Leona M. and Harry B. Helmsley Charitable Trust (Grant 2007-04026), Paul G. Allen Frontiers Group, and Stanford’s Plant Based Diet Initiative.

## 6. Author Contributions

DL and CK contributed to designing the study, interpreting the data and results, writing the manuscript, and preparing the figures. GH and CWP contributed to acquiring the data and performing data analysis. CT contributed to drafting and revising the manuscript. SG contributed to supervising the data acquisition and data analysis. SS contributed to supervising the study design, data acquisition, data analysis, manuscript writing and revision, and figure preparation. All authors reviewed the manuscript.

## 8. Data Availability Statement

The data that support the findings of this study are available from EVERSANA but restrictions apply to the availability of these data, which were used under license for the current study, and so are not publicly available. The authors will make a good faith effort to provide access to the data upon reasonable request.

