## Supplemental Table 1 for "Surveillance colonoscopy in patients with quiescent inflammatory bowel disease is associated with increased post-procedure steroid prescriptions: a national database study"

**Supplementary Data**

| **Cohort characteristic** | **Included codes** |
| --- | --- |
| IBD | ICD9:  [555] Regional enteritis  [556] Ulcerative colitis  ICD10:  [K50] Crohn disease (regional enteritis)  [K51] Ulcerative colitis |
| Colonoscopy | CPT:  [45395] Colonoscopy, flexible; with removal of tumor(s), polyp(s), or other lesion(s) by snare technique  [45390] Colonoscopy, flexible; with endoscopic mucosal resection |

**Table S1.** ICD9/10 and CPT codes used for selection of IBD cohort. ICD - International Classification of Diseases; CPT – Current Procedural Terminology.
